# Changes in EEG microstate dynamics and cognition post-chemotherapy in people with breast cancer: A pilot study

**DOI:** 10.1101/2024.07.03.24309730

**Authors:** S. Damji, S. Sattari, K. Zadravec, K.L. Campbell, J. Brunet, N. Virji-Babul

## Abstract

**Objective:** Chemotherapy-related cognitive changes following breast cancer are commonly reported; however, changes in brain dynamics of large-scale neural networks remain unclear. Using data from the *Aerobic exercise and CogniTIVe functioning in women with breAsT cancEr (ACTIVATE)* trial, we conducted exploratory analyses to compare self-reported and objective measures of cognition and applied microstate analysis to resting state electroencephalography (EEG) data of women with breast cancer before and following chemotherapy treatment.

**Methods:** Data from 8 female participants between the ages of 30 and 52 (mean age = 44.8 yrs, SD = 7.3 yrs) were analyzed. Cognitive function was assessed using the PROMIS (Patient-Reported Outcomes Measurement Information System) and the Trail Making Test (TMT). Five minutes of resting state, eyes-closed EEG data were also collected. Seven EEG microstates were extracted and mean microstate duration and occurrence were computed.

**Results:** Following chemotherapy, there was a significant decrease in the PROMIS score (*p* = 0.003, *d* = 1.601), but no significant difference in the TMT. Overall, durations of microstates were significantly longer (*p* < 0.001, *d* = 2.837) and less evenly distributed following chemotherapy. The mean duration of microstate D significantly increased following chemotherapy (*p* = 0.007, *d* = 1.339). No significant correlations between microstate features and the PROMIS score were observed.

**Conclusions:** We observed self-reported cognitive impairment and disturbed functional dynamics in the resting state brain following chemotherapy treatment. These results introduce a potential novel biomarker to evaluate the changes in large scale brain dynamics related to the cognitive effects of chemotherapy.

## INTRODUCTION

Chemotherapy-related cognitive impairment (CRCI), colloquially referred to as “chemo brain” or “chemo fog”, is a cluster of cognitive impairments including mental fatigue, attentional problems, and memory complaints^1–5^. Breast cancer survivors who have completed chemotherapy treatment report greater cognitive concerns post-treatment relative to pre-treatment, as well as greater executive dysfunction and impaired memory compared with matched healthy controls^6,7^. The neurophysiological correlates of CRCI remain unknown and currently there is no accepted “brain signature” of CRCI, making it difficult to develop strategies to manage this issue ^7^.

Standard electroencephalography (EEG) analysis methods have been applied to study CRCI in breast cancer survivors. Research primary focuses on isolated features of EEG, such as event-related potentials (ERP) and the static measurements of EEG band power underlying elevated physical and mental fatigue and changes in attentional processes^3,8,9^. However, these measures do not capture the dynamics of large scale brain networks.

Probing the brain during the resting state is a powerful way to map the functional organization of the brain, revealing that even when the brain is not involved in active cognitive tasks, it exhibits a highly organized pattern of activity characterized by synchronous neural activity over spatially distributed networks. These networks play a critical role in mediating complex functions such as memory, language, and emotional states^10,11^. EEG microstate analysis is a promising approach for studying the resting state of the brain by describing the discrete functional cortex-wide states that occur during rest. During the resting state, the brain does not exist in one state, but shifts dynamically between 4 to 7 different EEG “microstates” that are usually stable for about 20 to120 ms before shifting to another state^12–14^ and found to be highly reliable^15^. EEG microstates capture the subtle temporal dynamics in functional brain areas and networks that cannot be captured with resting-state fMRI alone, given the limited temporal resolution of the BOLD signal. Importantly, EEG microstates have strong associations with large-scale fMRI resting state brain networks (RSNs)^16,17^. Thus, this approach provides a novel lens for understanding altered dynamics of brain activity^18^.

In EEG microstate analysis, a modified k-means clustering algorithm is applied to segregate the multichannel time series data into fleeting epochs of cortex-wide electrical activity patterns (i.e., EEG microstates). Although 4 canonical microstate templates (A, B, C, D) have been most prevalent in the EEG microstate literature to date, Custo et al.^19^ argued that 7 distinct microstate templates best capture the scope of spontaneous electrophysiological activity topographies that are observed in resting-state EEG studies. Microstate A is associated with the auditory network; B with the visual network; C with the salience network; D with the attention network; E with the default mode network (DMN); F with cognitive and emotional processing related to activity in the anterior cingulate cortex (ACC); G with the sensorimotor network^19^. These microstates (labelled A-G) that have been reliably documented in the neuroscience literature and reproduced across numerous EEG studies^20^.

A growing number of studies have investigated EEG microstates in various clinical conditions to characterize the neural changes that occur in specific conditions with various degrees of success. For example, significant differences in microstate features have been reported in subjects with ALS as compared to healthy controls^21^. The dynamics of microstates C and D have also shown reliable differences in populations exhibiting psychotic symptoms: the brains of individuals affected by schizophrenia displayed a greater presence of microstate class C and a smaller presence of microstate class D as compared to healthy controls^16^. Interestingly, other mental health disturbances do not appear to manifest as aberrant EEG microstate features; a well-powered study showed that significant changes in EEG microstate mean duration and occurrence frequency were not observed in patients exhibiting mood and anxiety disorders^22^. Finally, preooperative microstate features were predictive of chronic postoperative pain perception in patients with breast cancer who underwent surgery^23^. To our knowledge, EEG microstate analysis has not yet been applied to studying CRCI in women with breast cancer.

An objective method to diagnose and monitor the severity of CRCI would be beneficial to patients with breast cancer and survivors. Such a method would not only provide women with valuable mechanistic insight into their symptoms, but also spur investigations into innovative interventions that may mitigate the impact of chemotherapy treatment on specific functional areas of the brain, as well as cognitive function in this cohort^24–26^. It has not been fully clarified if EEG can be used to track changes in the brain associated with CRCI, and in turn, the effects of potential interventions on these changes.

In this exploratory study, we investigate the potential of EEG microstates to provide a new electrophysiological perspective on functional activity changes related to chemotherapy treatment that can assist with the design of future clinical trials^27^. We also investigate the potential of EEG microstates as a biomarker for evaluating cognitive changes associated with CRCI. Given the previous literature showing changes in static measures of EEG activity associated with CRCI, we hypothesize that following chemotherapy treatment for breast cancer, individuals will show large scale changes in brain dynamics as measured by EEG microstates’ temporal features compared to before treatment.

## METHODS

### Participants

We analyzed data collected during 2018-2020 as part of a longitudinal randomized controlled trial entitled *Aerobic exercise and CogniTIVe functioning in women with breAsT cancEr (ACTIVATE)*^28^, which focused on testing the effects of aerobic exercise on cognition function and quality of life. In the ACTIVATE trial, participants were randomized to complete supervised, progressive, aerobic exercise during chemotherapy (EX) or wait-list usual care control (DE); in the present study, all participants were collapsed into a single group. See protocol for full details in Brunet et al.^28,29^. EEG data collection was optional for participants.

Ethics approval for the ACTIVATE trial was granted by the research ethics boards at the University of Ottawa (Ottawa, ON) and the University of British Columbia (UBC) (Vancouver; BC), as well as relevant hospital research ethics committees. The REB number for the data collected in Vancouver is H17-00563 (UBC/BC Cancer research ethics boards). This trial was registered with the ClinicalTrials.gov database (NCT03277898; September 11, 2017). All participants received written and oral information prior to participation and provide informed consent.

### Self-Reported and Objective Measures of Cognitive Function

The ACTIVATE trial included a wide range of outcome measures as described in Brunet et al.^28,29^. From these measures, we selected 1 self-report measure and 1 objective neuropsychological measure collected at baseline (before chemotherapy) and following chemotherapy to broadly capture the symptoms of cognitive impairment the sample experienced. For the self-report measure, we used the 4-item PROMIS (Patient-Reported Outcomes Measurement Information System) Applied Cognitive Abilities^30^ scale. For the objective cognitive function measure, we used the Trail Making Test (TMT), a standard assessment of cognitive flexibility, which is the ability to switch attentional resources between different tasks. The TMT consists of 2 separate scores for the time taken to complete 2 visual search tasks (i.e., part A and part B) and requires alternating attention in the most efficient manner possible. Higher scores (i.e., longer times) correspond to poorer cognitive flexibility.

### EEG Data Collection and Preprocessing

Baseline EEG testing was performed prior to starting chemotherapy (mean = 4.3 ± 6.1 days before the start of chemotherapy treatment). Post-chemotherapy EEG testing was a mean of 25.3 ± 18.4 days after treatment completion. Five-minute resting state EEG data were collected using 64-channel EGI HydroCel Geodesic SensorNets (EGI, Eugene, OR) amplified using Net Amps 300 with 500 Hz sampling rate and Cz as the reference. Scalp electrode impedances were kept bellow 50 *kΩ*.

Raw EEG data were preprocessed using EEGLAB (v2023.1)^31^ in MATLAB (v2023b). Each participant’s EEG was re-referenced to the average of all channels. We then applied a notch filter at 60 Hz, a low-pass filter at 50 Hz, and a high-pass filter at 0.5 Hz. Finally, Independent Component Analysis (ICA) was used to identify and remove any non-brain artifacts identified through a combination of visual inspection and EEGLAB’s ICLabel classification function^32^.

### EEG Microstate Analysis: Mean Duration and Mean Occurrence Frequency

In this exploratory study, we focused on 2 microstate features, namely microstate mean duration and frequency of occurrence, to determine if there were broad changes in the microstates between pre– and post-chemotherapy treatment. Mean duration reflects the temporal stability of a particular microstate, whereas frequency of occurrence represents the tendency of a particular microstate to be active (i.e., the frequency of usage of the functional brain network underlying a given microstate topography).

EEG microstate analysis was performed in MATLAB (v2023b) using the MICROSTATELAB toolbox (v1.0)^33^. Mean microstate maps were first generated for all participants at both timepoints (see Figure 2A) before being mapped onto the widely used microstate templates, and then backfitted (i.e., the raw EEG was re-expressed as a sequence of microstate classes) to each participants’ individual EEG time series for feature extraction. Following the advice of Nagabhushan Kalburgi et al.^33^, grand mean maps were used as the template for backfitting to ensure optimal comparability across participants and the most conservative analysis of extracted microstate features. At present, there is no consensus on how to determine the optimal number of classes to use for EEG microstate analysis^16^. Therefore, we sought a cluster solution that minimized globally explained variance (GEV) while also following the guidance that 7 microstate classes are needed to conduct the most comprehensive EEG microstate analysis of spontaneous electrophysiological activity in the brain^19^. The 7-class solution, as expected, yielded the highest GEV.

Backfitting was performed using 7 microstate classes on Global Field Power (GFP) peaks of the EEG recording. Microstate labels were interpolated in between these maxima using the nearest neighbour criterion, based on the most closely associated GFP label, as suggested by other researchers^19,26^. The mean duration and the mean frequency of occurrence per second were extracted for each microstate. Figure 1 summarizes this process.

**Figure 1.**
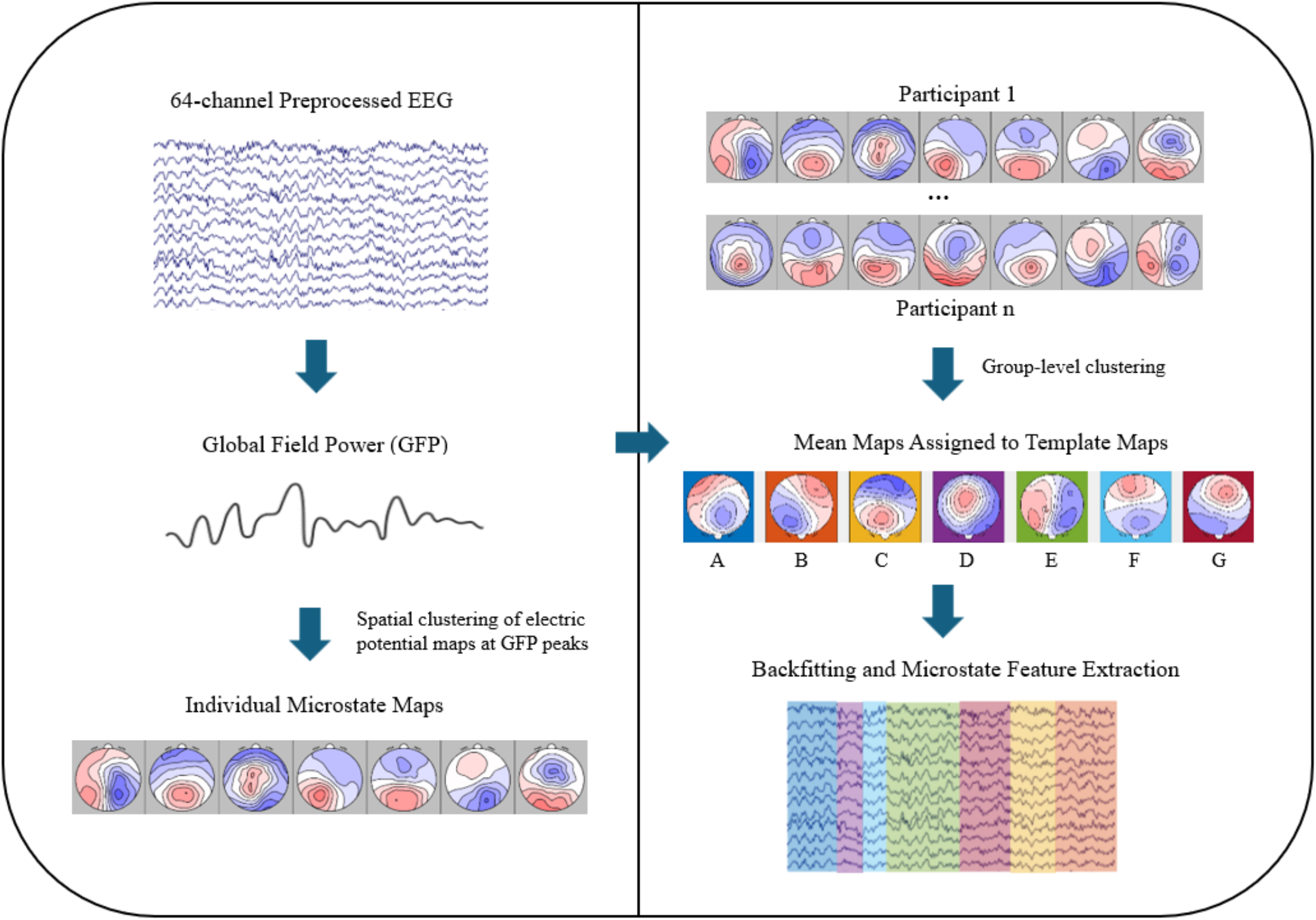
Flowchart outlining the microstate analysis process. Global Field Power (GFP) is first computed from an individual participant’s preprocessed EEG data. Next, electric potential topographical maps are derived at the GFP peaks. Individual-level spatial clustering using the k-means algorithm is performed on these topographical maps, yielding individual microstate maps. Following this, a second clustering process is applied across all individuals to generate group mean microstate maps. Mean maps are then sorted to normative template maps (A-G) based on shared variance. Finally, backfitting of these template maps at the subject level occurs, enabling the extraction of temporal features associated with individual microstates.

### Statistical Analysis

PROMS/TMT: Group-level paired t-tests were performed for the PROMIS score and both TMT scores to detect changes before and following chemotherapy treatment. Since 3 paired t-tests were performed, a Bonferroni-adjusted significance level of 0.017 was calculated to account for the increased probability of type-I error.

EEG paired sample t-tests were performed to evaluate statistically significant differences between both timepoints. The significance threshold was adjusted for multiple comparison; since we performed 7 comparisons (i.e., 1 for each microstate), we adjusted the significance level to 0.007.

Selected microstate features and measures of cognitive function that showed the largest differences between the baseline and post-chemotherapy timepoints were correlated to assess whether the most significant changes observed in the brain could be mapped onto the most significant indicators of cognitive impairment that were observed before versus after chemotherapy treatment.

## RESULTS

Eleven female participants diagnosed with stage I-III breast cancer completed a resting-state EEG scan and a battery of behavioral and neuropsychological tests before (i.e., baseline) and after (i.e., post-chemotherapy) the completion of chemotherapy treatment; due to 1 participant having EEG data that was too noisy for adequate preprocessing and analysis, and the attrition of an additional 2 participants at the post-chemotherapy timepoint, 8 participants (mean age = 44.8 yrs, SD = 7.3 yrs) were included in the final analysis for this study. One study participant had their baseline EEG scan performed after starting chemotherapy treatment. Their baseline EEG scan, however, was conducted just 1 week into their 24-week chemotherapy treatment protocol, and they had not yet completed the first cycle (lasting 3 weeks) of their chemotherapy treatment. 4 participants were in the EX group and 4 in the DE group. Table 1 summarizes the sociodemographic characteristics of the analytical sample.

**Table 1.** Demographic data and clinical characteristics of the cohort (n = 8). ^†^ One participant’s chemotherapy type could not be retrieved ^‡^ Doxorubicin, cyclophosphamide, paclitaxel ^§^ Agents in ACT + filgrastim ^¶^ Docetaxel, cyclophosphamide.

| Demographic/Clinical Variable | Group Summary |
| --- | --- |
| Age in years (mean ± SD) | 44.8 ± 7.3 |
| Ethnicity (Caucasian/Chinese) | 5/3 |
| Breast cancer stage (1/2/3) | 3/3/2 |
| Chemotherapy type<sup>†</sup> (ACT<sup>‡</sup>/ACT-G<sup>§</sup>/DC<sup>¶</sup>) | 4/1/2 |
| Chemotherapy length in weeks (mean ± SD) | 16.0 ± 3.7 |
| Patient-Reported Outcomes Measurement Information System Score (mean ± SD) | Baseline: 14.3 ± 5.0<br>Post-chemotherapy: 10.6 ± 4.6 |
| Trail Making Test Part A Score (mean ± SD) | Baseline: 31.5 ± 10.5<br>Post-chemotherapy: 26.9 ± 6.9 |
| Trail Making Test Part B Score (mean ± SD) | Baseline: 67.2 ± 22.3<br>Post-chemotherapy: 55.6 ± 12.0 |
| Days between baseline scan & chemotherapy start (mean ± SD) | 4.3 ± 6.1 |
| Days between post-chemotherapy scan & chemotherapy end (mean ± SD) | 25.3 ± 18.4 |

### Self-Reported and Objective Measures of Cognitive Function

Pre– and post-chemotherapy scores are reported in Table 1. At the Bonferroni-adjusted significance level, the PROMIS score significantly decreased following chemotherapy treatment and demonstrated a large effect size (*p* = 0.003, *d* = 1.601). Overall, participants self-reported significantly impaired memory, cognitive speed, and ability to focus on tasks following chemotherapy treatment.

Neither of the TMT scores following chemotherapy treatment were statistically significantly different from scores before treatment (part A: *p* = 0.283, *d* = 0.411; part B: *p* = 0.158, *d* = 0.558). Both scores decreased (i.e., corresponding to shorter times and thus better performance on the TMT) following chemotherapy treatment, with a small effect size observed for the time to complete part A, and a medium effect size observed for the time to complete part B. These effect sizes indicate that participants’ performance on the TMT showed a slight improvement following the chemotherapy treatment.

### EEG Microstate Results

Figure 2 shows the results for the global and individual microstate mean duration and mean occurrence at baseline and post-chemotherapy. Figure 2A shows the mean microstate maps generated for the cohort and organized based on the normative A to G maps.

**Figure 2.**
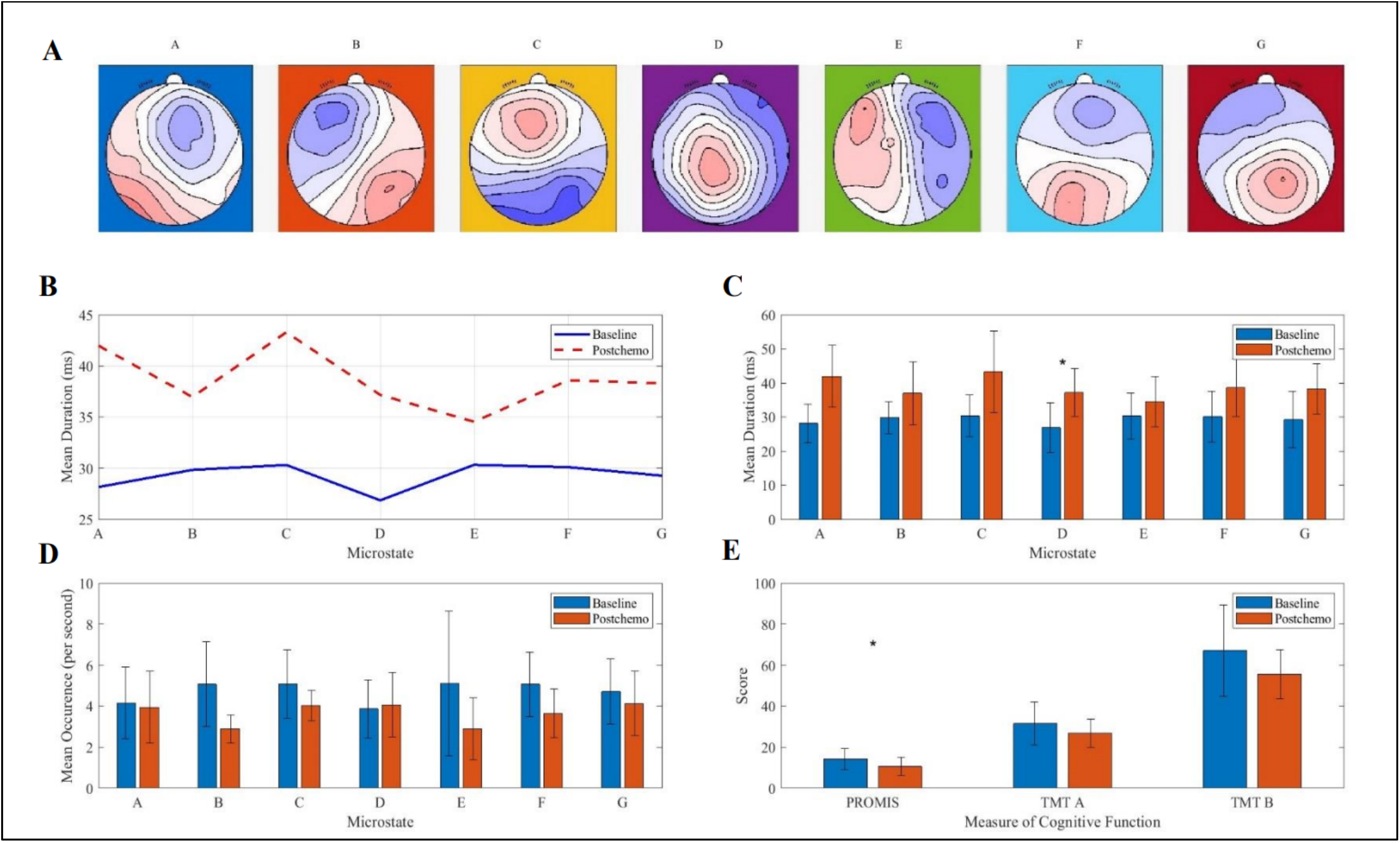
Overview of key results. **A** Mean microstate maps generated for our cohort and sorted to the normative A-G maps. **B** Overall, mean microstate durations following chemotherapy were significantly longer (*p* < 0.001, *d* = 2.837) and more than two times more variable (SD = 3.02) than mean microstate durations in the baseline group (SD = 1.31). **C** The mean duration of microstate D was significantly longer following chemotherapy (*p =* 0.007, *d =* 1.339). **D** There were no statistically significant results regarding the mean occurrence frequency of the microstates. **E** The PROMIS Cognitive Abilities total score was significantly reduced following chemotherapy treatment (*p* = 0.003, *d* = 1.601).

#### Microstates Mean Duration Observations

A paired t-test comparing the mean durations of all microstates before and after chemotherapy treatment showed that the mean durations significantly increased overall (Figure 2A), and this result exhibited a large effect size (*p* < 0.001, *d* = 2.837). The mean durations of the 7 microstates was also more consistent at baseline (ranging from 26.9 ms to 30.3 ms) compared to post-chemotherapy (ranging from 34.5 ms to 43.3 ms) based on the ranges observed.

Only the mean duration of microstate D showed a significant increase following chemotherapy *(p =* 0.007, *d =* 1.339). Based on observed large effect size, however, results of practical significance are the increase in mean duration of microstate A (*p =* 0.016, *d =* 1.120), microstate G (*p =* 0.043, *d =* 0.875), and microstate C (*p =* 0.044, *d =* 0.869).

#### Microstates Mean Occurrence Observations

At the Bonferroni-adjusted significance level of 0.007, none of comparisons of mean occurences for each of the 7 microstates between before and after chemotherapy treatment were statistically different. Based on the observed large effect size, however, results of practical significance are the reduction in the mean occurrence of microstate B (*p =* 0.018, *d =* 1.090) and microstate F (*p =* 0.029, *d =* 0.971). Moreover, the mean occurrence rate of microstates E (*p =* 0.177, *d=* 0.531) and C decreased (*p =* 0.256, *d=* 0.438) with a medium and small effect sizes, respectively. The mean occurrence rate of microstate D increased following chemotherapy but this result was of a negligible effect size and did not attain statistical significance *(p =* 0.765, *d =* 0.110).

#### Correlation Analysis

Six select microstate features were correlated with the PROMIS score at each timepoint. The microstate features selected included the mean durations of microstates A, C, D, and G, and the mean occurrences of microstates B and F; these particular microstate features showed the largest effect size between the baseline and post-chemotherapy timepoints. Using a Bonferroni-adjusted significance level of 0.008 to account for the increased probability of type-I error, none of the correlation tests reached statistical significance at either timepoint. Moreover, there were no visible differences in the magnitude of the correlation coefficients when comparing the baseline and post-chemotherapy timepoints.

## DISCUSSION

In this exploratory study, we investigated the changes in resting state EEG microstates in a small cohort of individuals with breast cancer followed from pre-to post-chemotherapy treatment and their possible associations with measures of cognitive function. The goal of this study was twofold: to explore a new method of analyzing the neural changes that occur as a result of chemotherapy treatment, and to generate specific hypotheses for future confirmatory and better-powered investigations. Overall, despite limitations due to small sample, notable changes were observed across each of the microstate features analyzed, as were changes in self-perceived cognitive function. These exploratory results support the conclusion that large-scale neural dynamics significantly changed in our cohort following chemotherapy, albeit these results should be subjected to additional examination in larger samples.

As expected based on prior research^1,34^, our sample of patients with breast cancer reported significantly greater cognitive impairments following 12 to 24 weeks of chemotherapy. These exploratory results provide further data to support calls to develop interventions to mitigate cognitive declines during treatment for breast cancer. Unexpectedly, however, participants’ scores on the TMT did not significantly change following chemotherapy; in fact, results indicated that their performance on this objective measure of cognitive function actually slightly improved following chemotherapy treatment. We suggest that it is unlikely that participants truly demonstrated improved cognitive flexibility following chemotherapy treatment, given participants’ own reports of their impaired cognitive capabilities—a subjective assessment that demonstrated a large effect size. It is possible that improvements in performance may have been due to familiarity with the TMT at timepoint two, cognitive reserve (i.e., the brain has the potential to actively compensate for cognitive impairment), increased effort (explained further), or an altered strategy.

Evidence for the impact of chemotherapy on the brain using EEG are sparse. In a healthy brain, individuals display a typical pattern of shifting between different brain states at rest, spending roughly equal amounts of time in each state^35^. In this cohort, we found that the overall durations of microstates were significantly longer and less evenly distributed following the completion of chemotherapy compared with the baseline status. We suggest that this finding may be related to the symptoms of “brain fog” which have been characterized as difficulty flexibly shifting between different cognitive states, such as shifting one’s attention from an internal state to paying attention to the external world^35^.

Our most significant finding was that the mean duration of microstate D increased following chemotherapy. Microstate D has been shown to primarily be associated with the attention, orientation, and executive function RSN (e.g., activity in the right inferior parietal lobe and the right middle and superior frontal gyri)^19,36^. Attention and executive functions are especially impaired in patients with breast cancer following chemotherapy treatment^3,7^. A disruption in these cognitive functions maps especially well onto symptoms of mental fatigue, difficulties with attention and memory impairment^6^. An increase in microstate D activation could be interpreted to reflect hyperactivity and a compensatory response response related to attention and executive function, likely associated with greater cognitive effort^6^. The increased effort and time in this state may also explain why we did not observe a significant difference in the scores of the TMT following chemotherapy treatment. Further study is needed to evaluate this finding.

Our result builds upon prior fMRI research^37^ and the review of fMRI studies conducted by Arya et al.^5^, showing that breast cancer treatment-related fatigue is associated with task-related frontoparietal hyperactivation (i.e., functional areas related to executive function), to possibly compensate for reduced activity in these regions at rest (i.e., reduced memory and planning abilities). Arya et al^5^, however, did not conduct a longitudinal analysis but rather a cross-sectional analysis of patients with breast cancer at various stages of treatment, relative to healthy controls and breast cancer patients not experiencing fatigue. Our finding suggests the possibility there is also within-subject heightened activity in these cortical areas at rest following chemotherapy treatment, relative to baseline.

Our findings with respect to microstates C and F were mixed. The mean duration for microstate C increased with a large effect size after chemotherapy, whereas we observed a small reduction in the mean occurrence of microstate C that can be attributed to our general observation that all microstate episodes were more prolonged following chemotherapy treatment.

Microstate C has been associated with the salience resting state network as well as functional areas associated with self-referential thought^36,38^. This finding adds a new perspective to the viewpoint that salience network connectivity changes can serve as an objective biomarker for cognitive impairments due to chemotherapy in breast cancer survivors^7^. Microstate F, on the other hand, demonstrated a large decrease in mean occurrence post-chemotherapy relative to baseline. There was, however, an unexpected increase in the mean duration of microstate F. This increase in mean duration reflects the general pattern of prolonged durations that we observed across all microstates. Microstate F has been associated with the activity of DMN^38^, and our finding that default mode activation may have been occurring less frequently post-chemotherapy aligns with the observation that several DMN connections may be disrupted following chemotherapy^6,39^.

Our preliminary results pertaining to microstates A, B, E, and G warrant further investigation. Seminal work by Britz et al.^36^ and Custo et al.^19^ has also showed that microstate A is primarily associated with the auditory processing RSN and language areas (e.g., the left middle and superior temporal lobe, including primary auditory cortex and the left Wernicke area) and that microstate B is primarily associated with the visual imagery RSN (e.g., activity in the left and right occipital cortices, including primary visual cortex). A more recent review by Tarailis et al.^38^, however, argued that microstate A may also be related to visual processing and arousal areas of the brain. Microstate topographies E and G have also been shown to be associated with various functional brain networks. Tarailis et al.^38^ argue that microstate E (moreso than microstate C) shows a strong association with the salience network, interoception, and the processing of autonomic information. Microstate G, on the other hand, may primarily be associated with activity in the somatosensory network^38^.

Our observed increase in the mean duration of microstate A but reduction in the mean occurrence of microstate B following chemotherapy (both results nearly attained significance and exhibited a large effect size) points to reduced activity in visual processing and imagery areas of the brain but increased activity in auditory and language processing areas of the brain following chemotherapy^19,36^. We also observed an increase in the mean duration of microstate G following chemotherapy, a microstate that has been associated with activity in the somatosensory network ^38^. Perhaps this altered balance of sensory processing activity in the brain, as a result of chemotherapy treatment, relates to the cognitive impairments that were experienced by our breast cancer cohort. Future research should further explore, as a primary research question, how chemotherapy treatment alters the processing of various sensory inputs in the brain, and whether or not an imbalance in brain activity related to the processing of different sensory inputs is a cause or effect of symptoms such as mental fatigue, attentional deficits, and memory impairment.

This exploratory study has key limitations. The small sample size and the number of statistical tests we performed (leading us to employ a conservative multiple testing correction) made statistical significance a less relevant outcome measure than measures of practical significance (i.e., standardized effect size). In addition, we did not differentiate between patients who received ACT (doxorubicin, cyclophosphamide, paclitaxel), patients who received ACT-G (agents in ACT + filgrastim), and patients who received DC (docetaxel, cyclophosphamide). A qEEG investigation by Vasaghi Gharamaleki^40^, however, suggests that different chemotherapy treatments may have different effects on the brains of patients with breast cancer. Other forms of cancer treatment, such as immunotherapy and radiotherapy, may also give rise to cognitive deficits that warrant further EEG microstate investigation ^27^. Finally, we analyzed data from a subset of participants from the ACTIVATE trial, where participants were randomized to complete supervised, progressive, aerobic exercise during chemotherapy or assigned to usual care, delayed exercise control condition. For our analysis, all participants were collapsed into a single group as the goal of this investigation was simply to observe differences before and after chemotherapy; still, the potential effect of undertaking aerobic exercise during chemotherapy may have impacted the findings.

In summary, this exploratory study provides evidence for self-reported cognitive impairment and disturbed functional dynamics in the resting state brain following chemotherapy treatment. These findings introduce EEG microstates as a potential novel biomarker to evaluate the changes in large scale brain dynamics related to the cognitive effects of chemotherapy, although future research is needed to confirm these exploratory findings. Additionally, whereas the present investigation focused on observing microstate dynamics before and after chemotherapy, future investigations may also want to explore the impact of various interventions during chemotherapy treatment aimed at attenuating or improving the experience of CRCI. Such interventions could include supplemental mindfulness-based exercises or aerobic exercise during the course of chemotherapy treatment^24^. Last, future research should parse out whether such interventions influence microstate outcomes and how these microstate outcomes may explain differences in cognitive function that are observed.

## Authors’ contributions

SD and SS conducted the analysis of the EEG data and the statistical analysis, drafted sections of the manuscript and critically revised and edited the manuscript. KZ assisted with the analysis of the behavioural data and critically reviewed and edited the manuscript. KLC and JB conceptualized the ACTIVATE trial, acquired funding, oversaw the conduct of the trial and critically reviewed and edited the manuscript. NVB contributed to the EEG trial design and critically reviewed and edited the manuscript. All authors read and approved the final manuscript.

## Acknowledgements

The authors would like to thank all the participants who took part in this study.

## Data availability

The raw data supporting the conclusions of this manuscript cannot be made available by the authors as patients were assured their data would be kept private and confidential to the extent permitted by law and that only the research team would have access to the data.

